# Impact of disasters, including pandemics, on cardiometabolic outcomes across the life-course: A systematic review

**DOI:** 10.1101/2020.11.27.20239830

**Authors:** Vanessa De Rubeis, Jinhee Lee, Muhammad Saqib Anwer, Yulika Yoshida-Montezuma, Alessandra T. Andreacchi, Erica Stone, Saman Iftikhar, Jason D. Morgenstern, Reid Rebinsky, Sarah E. Neil-Sztramko, Elizabeth Alvarez, Emma Apatu, Laura N. Anderson

## Abstract

**Objectives:** Disasters, such as the current COVID-19 pandemic, disrupt daily life, increase uncertainty and stress, and may increase long-term risk of adverse cardiometabolic outcomes, including heart disease, obesity and diabetes. The objective was to conduct a systematic review to determine the impact of disasters, including pandemics, on cardiometabolic outcomes across the life-course.

**Design:** A systematic search was conducted in May 2020 using two electronic databases, EMBASE and Medline. All studies were screened in duplicate at title and abstract, and full-text level. Studies were eligible for inclusion if they assessed an association with population-level or community disaster and cardiometabolic outcomes. There were no restrictions on year of publication, country or population. Non-English and earthquake-related studies were excluded. Data were extracted on study characteristics, exposure (e.g., type of disaster, name of specific event, region, year), cardiometabolic outcomes, and measures of effect. Study quality was evaluated using the Joanna Briggs Institute critical appraisal tools.

**Results:** A total of 58 studies were included, with 24 studies reporting the effects of exposure to disaster during pregnancy/childhood and 34 studies reporting the effects of exposure during adulthood. Studies included exposure to natural (60%) and human-made (40%) disasters, with only 3 (5%) of these studies evaluating previous pandemics. Most studies were conducted in North America (62%). Most studies reported increased cardiometabolic risk, including increased cardiovascular disease incidence or mortality, diabetes, and obesity. Few studies investigated potential mechanisms or identified high risk subgroups.

**Conclusions:** Understanding the long-term consequences of disasters on cardiometabolic outcomes across the life-course may inform public health strategies for the current COVID-19 pandemic. This review found strong evidence of an increased association between disaster exposure and cardiometabolic outcomes across the life-course, although more research is needed to better understand the mechanisms and preventative efforts.

**PROSPERO registration:** CRD – 42020186074

**Strengths and limitations of this study:**

- This systematic review is one of the first to review the literature on disasters, including pandemics, and subsequent cardiometabolic outcomes throughout the life-course.
- A comprehensive search strategy was developed in consultation with Health Science Librarians at McMaster University, which resulted in 58 studies that were eligible for inclusion into the review.
- Due to the heterogeneity of the included studies, a meta-analysis was not conducted.
- This review contributes a synthesis of the literature on the impact of disasters and cardiometabolic outcomes, that can help to inform public health strategies for the current COVID-19 pandemic.

## BACKGROUND

Disasters as defined by the World Health Organization (WHO), are events that disrupt the daily functioning of a community or society causing material, economic or environmental losses, overwhelming local capacity (1). Disasters can be categorized into natural disasters, human-made disasters, and hybrid disasters (2). Natural disasters include natural phenomenon above and beneath the earths surface (e.g., tsunamis or landslides), meteorological phenomenon (e.g., tornadoes or floods) or biological phenomenon (e.g., epidemics and pandemics) (2). Human-made disasters include adverse transportation incidents, technological events (e.g., fire or toxic leaks), terrorism, warfare or conflict (2). A hybrid disaster results from both human error and natural forces, such as the clearing of a jungle that results in a landslide (2). All types of disasters can result in public health emergencies as they typically impact a significant proportion of people (3). Epidemics, defined as a greater than expected increase in cases of a disease, and pandemics, which cross countries and continents, are types of natural disasters with far-reaching global disruption (4). The COVID-19 pandemic is a present-day example of a global disaster that is unlike any disaster in recent history. Understanding the potential long-term health implications of the current COVID-19 pandemic and resulting public health mitigation strategies is urgently needed.

Previous systematic reviews have focused specifically on the psychological impact of quarantine during pandemics (5), the impact on health outcomes after disasters in older adults (6), medically unexplained physical symptoms following disasters (7), and chronic medical interventions following a natural disaster (8). However, there is currently no review specifically focusing on the impact of either disasters, or epidemics and pandemics on cardiometabolic outcomes across the life-course. Noncommunicable diseases (NCDs), including cardiovascular disease (CVD), obesity, and diabetes, are the leading cause of morbidity and mortality worldwide (9,10). NCDs are attributed to 71% of all global deaths annually, with approximately 14 million CVD-related deaths and 1.6 million diabetes-related deaths (10). Findings from the Global Burden of Diseases Study indicate that CVD and diabetes account for over 20% of the global burden of disability, with diabetes recently emerging as the fourth leading cause of disability globally (9). Exposure to disasters may cause cardiometabolic outcomes to emerge or worsen through several different mechanistic pathways including stress exposure (11), lack of access to health services (12), food security and behavioural changes such as alterations in physical activity, sleep, and diet (13). It is important to understand the impact of disasters on the incidence of new cardiometabolic diseases and changes in disease status in all populations and age groups. Particular subgroups of a population may be more or less susceptible to cardiometabolic outcomes and understanding this can inform targeted interventions. The primary objective of this review was to determine the impact of disasters, including pandemics such as COVID-19, on risk of cardiometabolic outcomes across the life-course. The secondary objectives were to determine how to reduce the impact of chronic disease outcomes following a disaster and to identify populations at highest risk of cardiometabolic outcomes following a disaster.

## METHODS

A systematic review was conducted following the Preferred Reporting Items for Systematic Reviews and Meta-Analyses (PRISMA) (14). This review was registered on PROSPERO (CRD – 42020186074).

### Search strategy

A systematic search was conducted in May 2020 using the electronic databases EMBASE and MEDLINE. The health research librarians at McMaster University assisted in developing the search strategy. The search broadly captured two concepts: disasters and cardiometabolic outcomes (e.g., diabetes, obesity, hypertension). The complete search strategy for EMBASE can be found in Table 1. The search strategy for MEDLINE can be found in the Appendix (Table A1). Reference lists of eligible studies and relevant systematic reviews were hand searched to identify additional articles.

**Table 1.** Search strategy for EMBASE

|  |  |  |
| --- | --- | --- |
| 1 | social isolation.mp. or social isolation/ | 24963 |
| 2 | quarantine.mp. or quarantine/ | 4752 |
| 3 | *epidemic/ | 32686 |
| 4 | *pandemic/ | 4387 |
| 5 | disease outbreak.mp. | 2321 |
| 6 | disaster/ | 13321 |
| 7 | *natural disaster/ | 968 |
| 8 | humanitarian crisis.mp. | 257 |
| 9 | mass casualty.mp. or mass disaster/ | 3654 |
| 10 | coronavirus.mp. or coronaviridae/ | 23106 |
| 11 | cardiovascular disease.mp. or *cardiovascular disease/ | 357319 |
| 12 | *diabetes mellitus/ | 210248 |
| 13 | *cerebrovascular accident/ | 78444 |
| 14 | *heart infarction/ | 99072 |
| 15 | *angina pectoris/ | 22631 |
| 16 | *obesity/ | 178134 |
| 17 | public health emergency.mp. | 1752 |
| 18 | *body mass/ | 31459 |
| 19 | *hypertension/ | 198593 |
| 20 | 1 or 2 or 3 or 4 or 5 or 6 or 7 or 8 or 9 or 10 or 17 | 109105 |
| 21 | 11 or 12 or 13 or 14 or 15 or 16 or 18 or 19 | 1087681 |
| 22 | 20 and 21 | 2047 |
| 23 | limit 22 to human | 1832 |

### Eligibility criteria

Studies were eligible for inclusion if they assessed the relationship between a population-level or community disaster and the risk of future cardiometabolic outcomes including CVD, diabetes or obesity or cardiometabolic risk scores (15). CVD included myocardial infarction, stroke, hypertension and angina. There were no restrictions on year of publication, country of disaster, or population. Only studies evaluating the impact of real-world disasters in humans were included. Due to the research team’s capacity, only studies published in English were included. Observational and quasi-experimental study designs, including case-control, cohort and other longitudinal study designs or natural experiments were included. Outcomes that were not cardiometabolic-related or acute cardiometabolic events, such as an immediate complication (defined as <1 month after disaster), were excluded. Studies that assessed the exposure to a chemical as a result of the disaster, were excluded, as we were not interested in outcomes resulting from chemical exposure. Earthquake studies were also excluded since a systematic review was published in 2018 that assessed the impact of earthquakes on cardiometabolic outcomes (16).

### Study selection

After running the search, all identified studies were imported into Covidence and duplicates were removed (17). Studies were screened at title and abstract-level, and then at full-text by two independent reviewers (VD, JL, MSA, YYM, ATA, ES, SI, JDM, RR LNA). Conflicts were resolved by a third reviewer who made the final decision regarding eligibility for inclusion.

### Data extraction

A data extraction template was created, and pilot tested prior to data extraction. Data were then extracted from all studies by two independent abstractors (VD, JL, SMA, YYM, ATA, RR) and conflicts were resolved by a third independent abstractor. Study characteristics including year of publication, study design, country of disaster, sample size, and length of study were extracted where reported. Specific information on the exposure and outcome in each study were extracted including the type and name of the disaster, country and year of the disaster, the outcome of interest, and how the exposure and outcome were measured. Finally, any information on subgroups including age, population, sex and disaster type were also extracted, if applicable.

### Critical appraisal

Critical appraisal was conducted using the Joanna Briggs Institute (JBI) Critical Appraisal Tools (18). This tool was chosen due to the availability of checklists for a wide range of study designs, including cohort, cross-sectional and quasi-experimental designs (18). The quasi-experimental study design checklist was used for natural experiments including time-series studies and pre/post designs, as it was decided this was the most appropriate tool. All studies were critically appraised independently by two individuals (VD, JL, MA, YYM, ATA, ES, SI) and a third individual was consulted for any discrepancies.

### Data analysis

Data from the included studies were narratively synthesized. Results are presented by exposure period (perinatal/childhood versus adulthood) and by cardiometabolic outcome (obesity, CVD, and diabetes). Characteristics of studies are presented as frequencies and percentages. Due to the heterogeneity of studies, a meta-analysis was not conducted.

## RESULTS

A total of 4830 studies were identified through the electronic database search. An additional 12 studies were identified through manual searching of the reference lists of relevant studies. After removing duplicates (n=439), 4403 studies were screened at title and abstract level. After applying inclusion and exclusion criteria, 4068 studies were excluded, leaving 335 studies screened for full-text eligibility. A total of 58 studies were eligible for inclusion into the review. The complete screening process is described in Figure 1.

**Figure 1.**
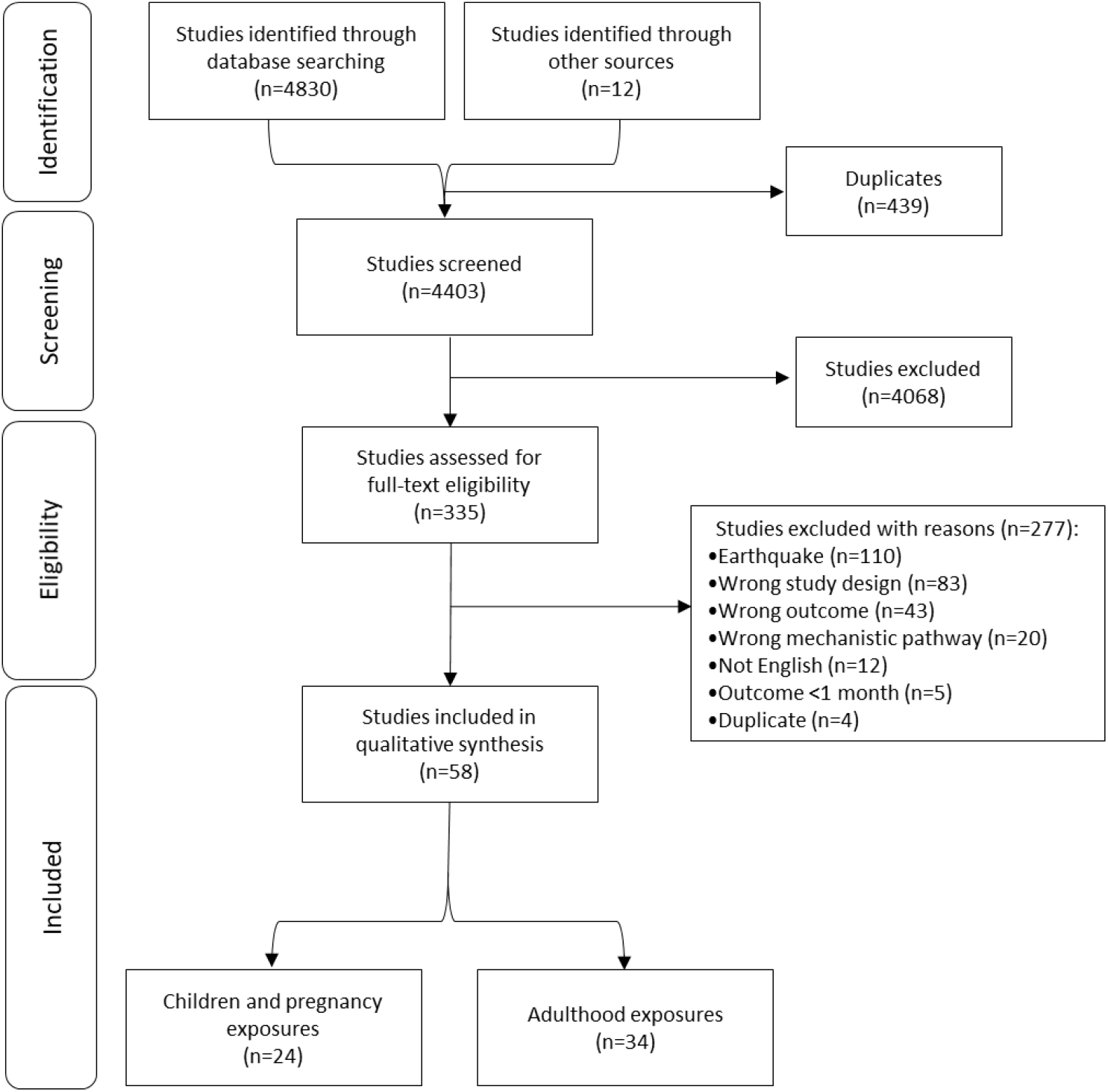
PRISMA Flow Chart.

### Description of studies

Table 2 provides a summary of included studies. Of 58 included studies, 24 studies (13,19–41) investigated exposure to disasters during pregnancy or childhood while the remaining 34 studies (12,42–74) investigated exposure to disaster during adulthood. Almost all studies (n=49) assessed cardiometabolic outcomes during adulthood, only two studies assessed outcomes during pregnancy (25,26) and seven studies assessed outcomes during childhood and adolescence (13,19–24). The length of studies, including prospective follow-up and retrospective assessment, ranged from 1 month to 95 years. Most studies (n=36) focused on disasters that occurred in North America (12,19–26,36,37,40,43–47,49,50,52,53,55–58,61–63,65–70,73,74), followed by Europe (n=13) (27–33,39,41,51,60,71,72). The remaining disasters occurred in Asia (n=7) (13,35,38,42,48,54,59) and Africa (n=2) (34,64). The characteristics of included studies and key findings are provided in Table 3 for disaster exposure during the perinatal period and childhood, and Table 4 for exposure during adulthood.

**Table 2.**
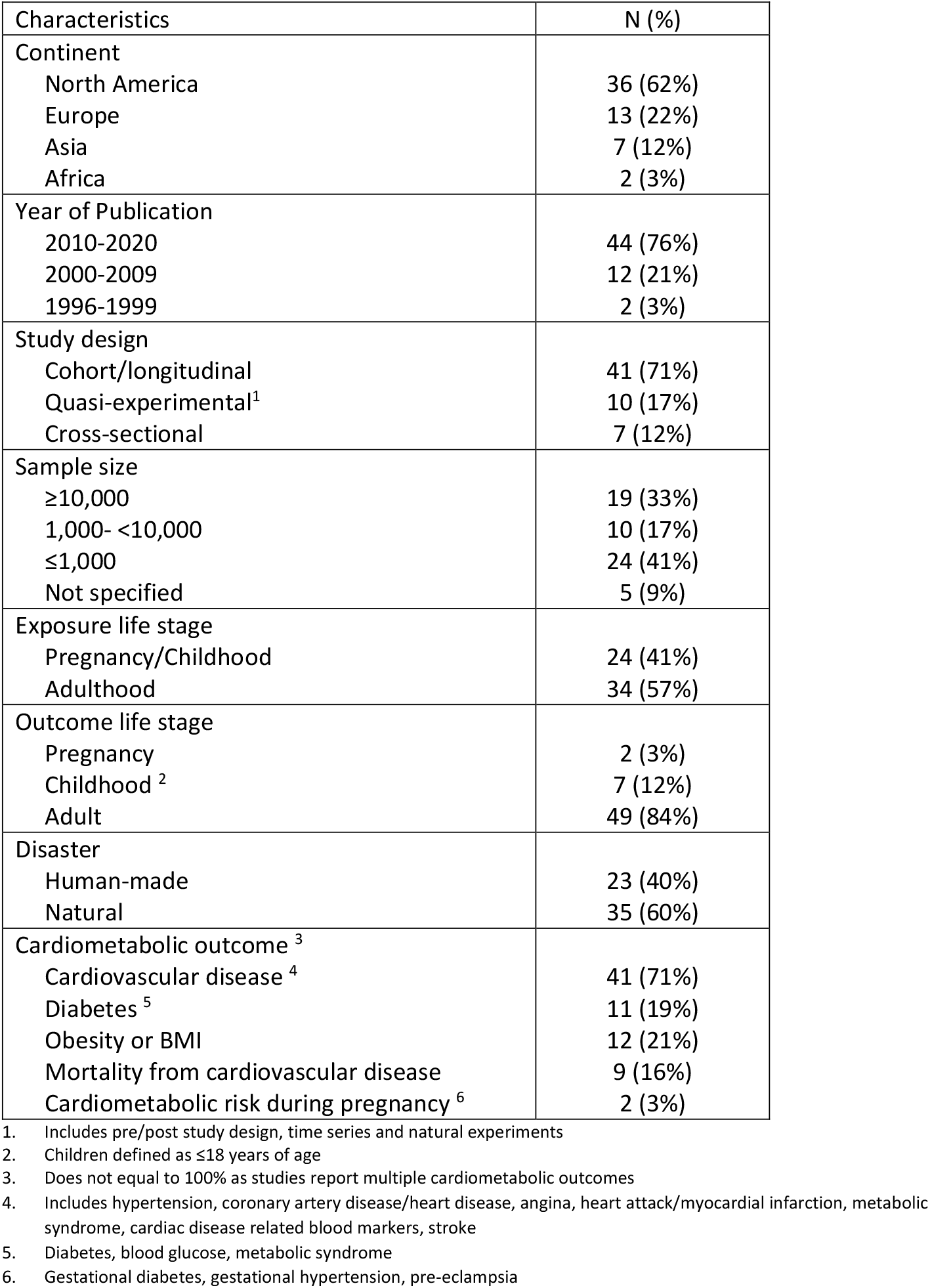
Characteristics of included studies (n=58)

**Table 3.**
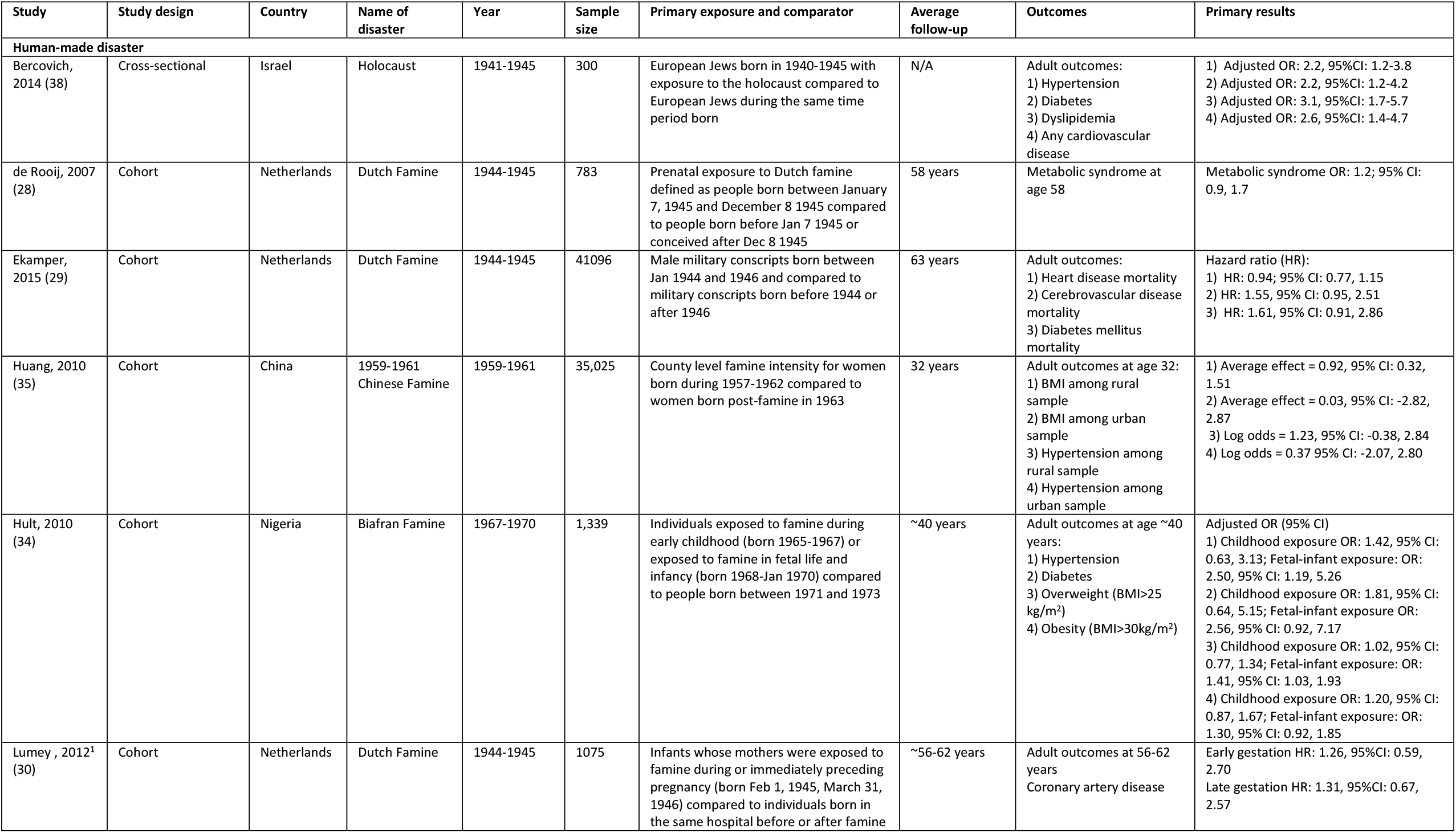
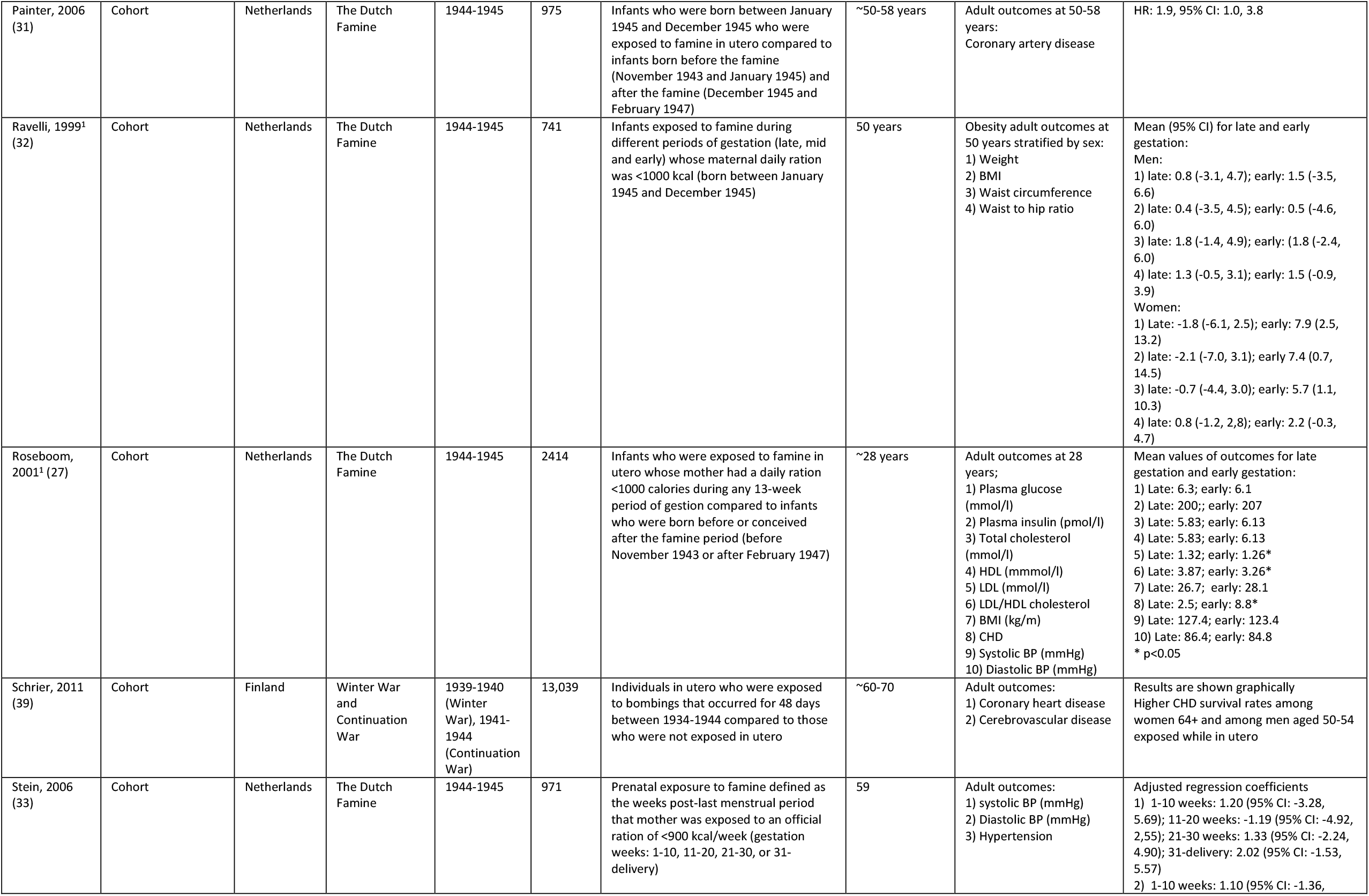
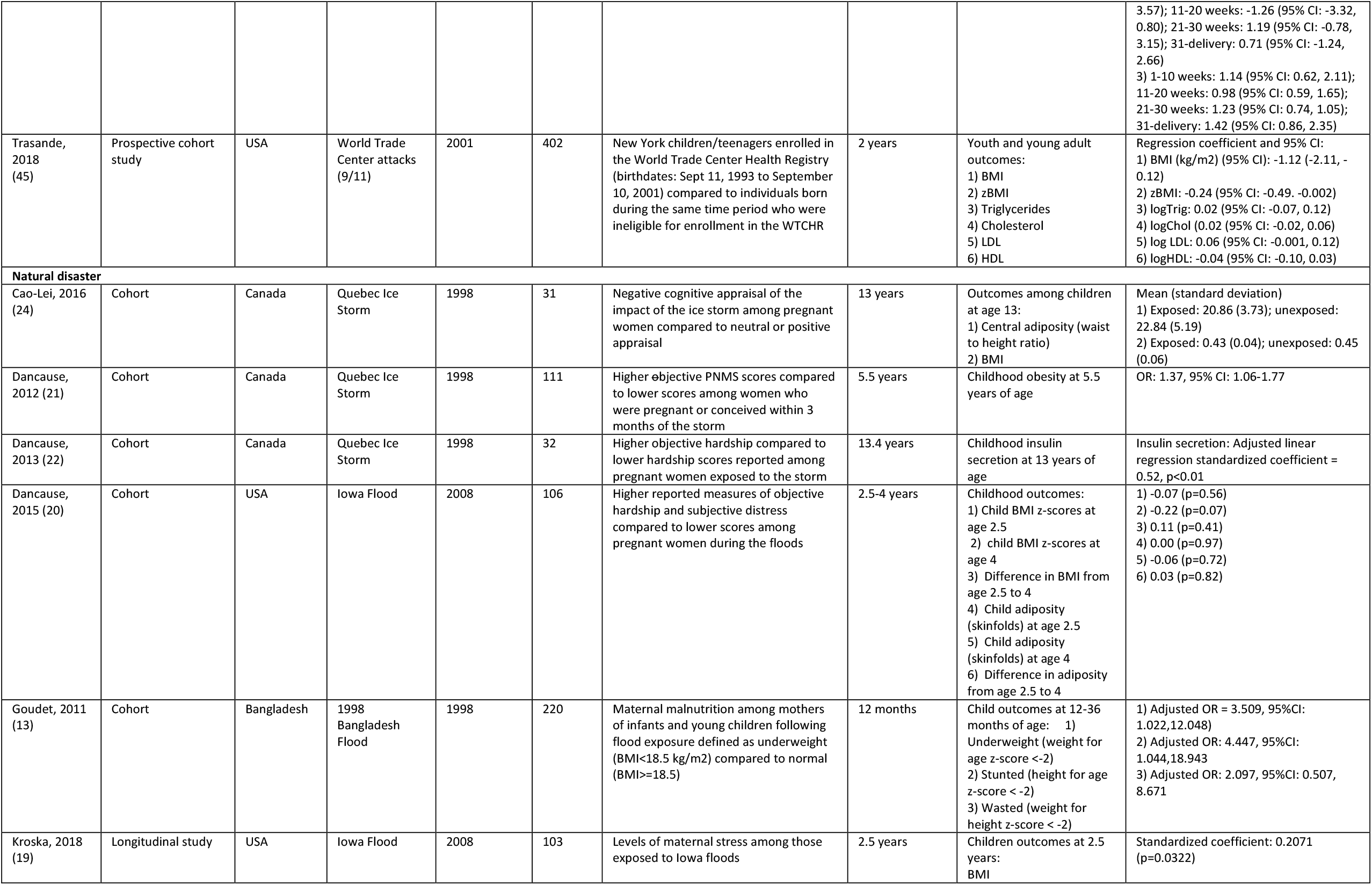
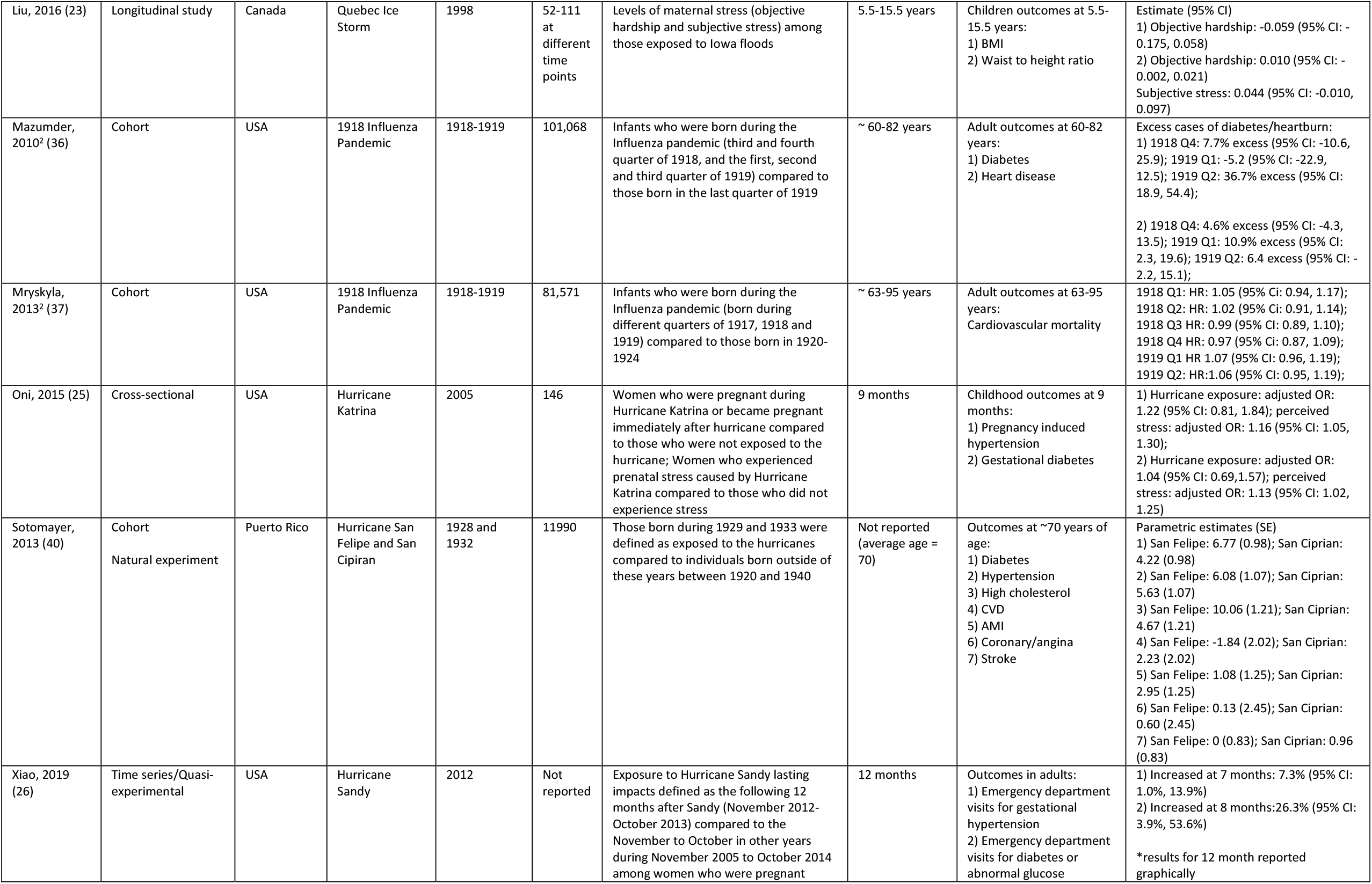
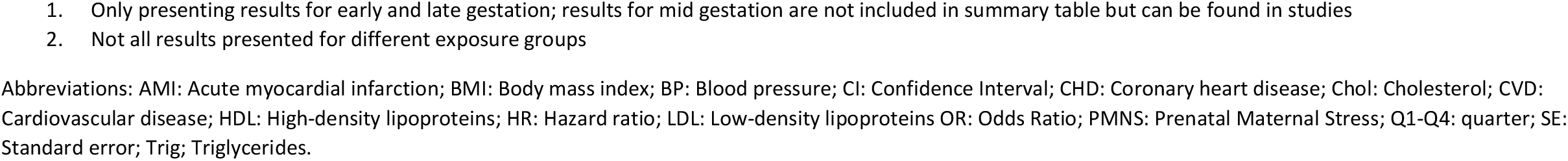
Characteristics of included studies investigating the association between exposure to a disaster during the perinatal and childhood periods and cardiometabolic outcomes across the life-course, by disaster type (n=24)

| Study | Study design | Country | Name of disaster | Year | Sample size | Primary exposure and comparator | Average follow-up | Outcomes | Primary results |
| --- | --- | --- | --- | --- | --- | --- | --- | --- | --- |
| **Human-made disaster** |  |  |  |  |  |  |  |  |  |
| Bercovich, 2014 (38) | Cross-sectional | Israel | Holocaust | 1941-1945 | 300 | European Jews born in 1940-1945 with exposure to the holocaust compared to European Jews during the same time period born | N/A | Adult outcomes:<br>1) Hypertension<br>2) Diabetes<br>3) Dyslipidemia<br>4) Any cardiovascular disease | 1) Adjusted OR: 2.2, 95%CI: 1.2-3.8<br>2) Adjusted OR: 2.2, 95%CI: 1.2-4.2<br>3) Adjusted OR: 3.1, 95%CI: 1.7-5.7<br>4) Adjusted OR: 2.6, 95%CI: 1.4-4.7 |
| de Rooij, 2007 (28) | Cohort | Netherlands | Dutch Famine | 1944-1945 | 783 | Prenatal exposure to Dutch famine defined as people born between January 7, 1945 and December 8 1945 compared to people born before Jan 7 1945 or conceived after Dec 8 1945 | 58 years | Metabolic syndrome at age 58 | Metabolic syndrome OR: 1.2; 95% CI: 0.9, 1.7 |
| Ekamper, 2015 (29) | Cohort | Netherlands | Dutch Famine | 1944-1945 | 41096 | Male military conscripts born between Jan 1944 and 1946 and compared to military conscripts born before 1944 or after 1946 | 63 years | Adult outcomes:<br>1) Heart disease mortality<br>2) Cerebrovascular disease mortality<br>3) Diabetes mellitus mortality | Hazard ratio (HR):<br>1) HR: 0.94; 95% CI: 0.77, 1.15<br>2) HR: 1.55, 95% CI: 0.95, 2.51<br>3) HR: 1.61, 95% CI: 0.91, 2.86 |
| Huang, 2010 (35) | Cohort | China | 1959-1961 Chinese Famine | 1959-1961 | 35,025 | County level famine intensity for women born during 1957-1962 compared to women born post-famine in 1963 | 32 years | Adult outcomes at age 32:<br>1) BMI among rural sample<br>2) BMI among urban sample<br>3) Hypertension among rural sample<br>4) Hypertension among urban sample | 1) Average effect = 0.92, 95% CI: 0.32, 1.51<br>2) Average effect = 0.03, 95% CI: -2.82, 2.87<br>3) Log odds = 1.23, 95% CI: -0.38, 2.84<br>4) Log odds = 0.37 95% CI: -2.07, 2.80 |
| Hult, 2010 (34) | Cohort | Nigeria | Biafran Famine | 1967-1970 | 1,339 | Individuals exposed to famine during early childhood (born 1965-1967) or exposed to famine in fetal life and infancy (born 1968-Jan 1970) compared to people born between 1971 and 1973 | ~40 years | Adult outcomes at age ~40 years:<br>1) Hypertension<br>2) Diabetes<br>3) Overweight (BMI>25 kg/m<sup>2</sup>)<br>4) Obesity (BMI>30kg/m<sup>2</sup>) | Adjusted OR (95% CI)<br>1) Childhood exposure OR: 1.42, 95% CI: 0.63, 3.13; Fetal-infant exposure OR: 2.50, 95% CI: 1.19, 5.26<br>2) Childhood exposure OR: 1.81, 95% CI: 0.64, 5.15; Fetal-infant exposure OR: 2.56, 95% CI: 0.92, 7.17<br>3) Childhood exposure OR: 1.02, 95% CI: 0.77, 1.34; Fetal-infant exposure OR: 1.41, 95% CI: 1.03, 1.93<br>4) Childhood exposure OR: 1.20, 95% CI: 0.87, 1.67; Fetal-infant exposure OR: 1.30, 95% CI: 0.92, 1.85 |
| Lumey, 2012<sup>1</sup> (30) | Cohort | Netherlands | Dutch Famine | 1944-1945 | 1075 | Infants whose mothers were exposed to famine during or immediately preceding pregnancy (born Feb 1, 1945, March 31, 1946) compared to individuals born in the same hospital before or after famine | ~56-62 years | Adult outcomes at 56-62 years<br>Coronary artery disease | Early gestation HR: 1.26, 95%CI: 0.59, 2.70<br>Late gestation HR: 1.31, 95%CI: 0.67, 2.57 |
| Painter, 2006 (31) | Cohort | Netherlands | The Dutch Famine | 1944-1945 | 975 | Infants who were born between January 1945 and December 1945 who were exposed to famine in utero compared to infants born before the famine (November 1943 and January 1945) and after the famine (December 1945 and February 1947) | ~50-58 years | Adult outcomes at 50-58 years:<br>Coronary artery disease | HR: 1.9, 95% CI: 1.0, 3.8 |
| Ravelli, 1999<sup>1</sup> (32) | Cohort | Netherlands | The Dutch Famine | 1944-1945 | 741 | Infants exposed to famine during different periods of gestation (late, mid and early) whose maternal daily ration was <1000 kcal (born between January 1945 and December 1945) | 50 years | Obesity adult outcomes at 50 years stratified by sex:<br>1) Weight<br>2) BMI<br>3) Waist circumference<br>4) Waist to hip ratio | Mean (95% CI) for late and early gestation:<br>Men:<br>1) late: 0.8 (-3.1, 4.7); early: 1.5 (-3.5, 6.6)<br>2) late: 0.4 (-3.5, 4.5); early: 0.5 (-4.6, 6.0)<br>3) late: 1.8 (-1.4, 4.9); early: (1.8 (-2.4, 6.0)<br>4) late: 1.3 (-0.5, 3.1); early: 1.5 (-0.9, 3.9)<br>Women:<br>1) Late: -1.8 (-6.1, 2.5); early: 7.9 (2.5, 13.2)<br>2) late: -2.1 (-7.0, 3.1); early 7.4 (0.7, 14.5)<br>3) late: -0.7 (-4.4, 3.0); early: 5.7 (1.1, 10.3)<br>4) late: 0.8 (-1.2, 2.8); early: 2.2 (-0.3, 4.7) |
| Roseboom, 2001<sup>1</sup> (27) | Cohort | Netherlands | The Dutch Famine | 1944-1945 | 2414 | Infants who were exposed to famine in utero whose mother had a daily ration <1000 calories during any 13-week period of gestation compared to infants who were born before or conceived after the famine period (before November 1943 or after February 1947) | ~28 years | Adult outcomes at 28 years;<br>1) Plasma glucose (mmol/l)<br>2) Plasma insulin (pmol/l)<br>3) Total cholesterol (mmol/l)<br>4) HDL (mmol/l)<br>5) LDL (mmol/l)<br>6) LDL/HDL cholesterol<br>7) BMI (kg/m)<br>8) CHD<br>9) Systolic BP (mmHg)<br>10) Diastolic BP (mmHg) | Mean values of outcomes for late gestation and early gestation:<br>1) Late: 6.3; early: 6.1<br>2) Late: 200; early: 207<br>3) Late: 5.83; early: 6.13<br>4) Late: 5.83; early: 6.13<br>5) Late: 1.32; early: 1.26*<br>6) Late: 3.87; early: 3.26*<br>7) Late: 26.7; early: 28.1<br>8) Late: 2.5; early: 8.8*<br>9) Late: 127.4; early: 123.4<br>10) Late: 86.4; early: 84.8<br>* p<0.05 |
| Schrier, 2011 (39) | Cohort | Finland | Winter War and Continuation War | 1939-1940 (Winter War), 1941-1944 (Continuation War) | 13,039 | Individuals in utero who were exposed to bombings that occurred for 48 days between 1934-1944 compared to those who were not exposed in utero | ~60-70 | Adult outcomes:<br>1) Coronary heart disease<br>2) Cerebrovascular disease | Results are shown graphically<br>Higher CHD survival rates among women 64+ and among men aged 50-54 exposed while in utero |
| Stein, 2006 (33) | Cohort | Netherlands | The Dutch Famine | 1944-1945 | 971 | Prenatal exposure to famine defined as the weeks post-last menstrual period that mother was exposed to an official ration of <900 kcal/week (gestation weeks: 1-10, 11-20, 21-30, or 31-delivery) | 59 | Adult outcomes:<br>1) systolic BP (mmHg)<br>2) Diastolic BP (mmHg)<br>3) Hypertension | Adjusted regression coefficients<br>1) 1-10 weeks: 1.20 (95% CI: -3.28, 5.69); 11-20 weeks: -1.19 (95% CI: -4.92, 2.55); 21-30 weeks: 1.33 (95% CI: -2.24, 4.90); 31-delivery: 2.02 (95% CI: -1.53, 5.57)<br>2) 1-10 weeks: 1.10 (95% CI: -1.36, |
|  |  |  |  |  |  |  |  |  | 3.57); 11-20 weeks: -1.26 (95% CI: -3.32, 0.80); 21-30 weeks: 1.19 (95% CI: -0.78, 3.15); 31-delivery: 0.71 (95% CI: -1.24, 2.66)<br>3) 1-10 weeks: 1.14 (95% CI: 0.62, 2.11); 11-20 weeks: 0.98 (95% CI: 0.59, 1.65); 21-30 weeks: 1.23 (95% CI: 0.74, 1.05); 31-delivery: 1.42 (95% CI: 0.86, 2.35) |
| Trasande, 2018 (45) | Prospective cohort study | USA | World Trade Center attacks (9/11) | 2001 | 402 | New York children/teenagers enrolled in the World Trade Center Health Registry (birthdates: Sept 11, 1993 to September 10, 2001) compared to individuals born during the same time period who were ineligible for enrollment in the WTCRH | 2 years | Youth and young adult outcomes:<br>1) BMI<br>2) zBMI<br>3) Triglycerides<br>4) Cholesterol<br>5) LDL<br>6) HDL | Regression coefficient and 95% CI:<br>1) BMI (kg/m2) (95% CI): -1.12 (-2.11, -0.12)<br>2) zBMI: -0.24 (95% CI: -0.49, -0.002)<br>3) logTrig: 0.02 (95% CI: -0.07, 0.12)<br>4) logChol (0.02 (95% CI: -0.02, 0.06)<br>5) log LDL: 0.06 (95% CI: -0.001, 0.12)<br>6) logHDL: -0.04 (95% CI: -0.10, 0.03) |
| **Natural disaster** |  |  |  |  |  |  |  |  |  |
| Cao-Lei, 2016 (24) | Cohort | Canada | Quebec Ice Storm | 1998 | 31 | Negative cognitive appraisal of the impact of the ice storm among pregnant women compared to neutral or positive appraisal | 13 years | Outcomes among children at age 13:<br>1) Central adiposity (waist to height ratio)<br>2) BMI | Mean (standard deviation)<br>1) Exposed: 20.86 (3.73); unexposed: 22.84 (5.19)<br>2) Exposed: 0.43 (0.04); unexposed: 0.45 (0.06) |
| Dancause, 2012 (21) | Cohort | Canada | Quebec Ice Storm | 1998 | 111 | Higher objective PNMS scores compared to lower scores among women who were pregnant or conceived within 3 months of the storm | 5.5 years | Childhood obesity at 5.5 years of age | OR: 1.37, 95% CI: 1.06-1.77 |
| Dancause, 2013 (22) | Cohort | Canada | Quebec Ice Storm | 1998 | 32 | Higher objective hardship compared to lower hardship scores reported among pregnant women exposed to the storm | 13.4 years | Childhood insulin secretion at 13 years of age | Insulin secretion: Adjusted linear regression standardized coefficient = 0.52, p<0.01 |
| Dancause, 2015 (20) | Cohort | USA | Iowa Flood | 2008 | 106 | Higher reported measures of objective hardship and subjective distress compared to lower scores among pregnant women during the floods | 2.5-4 years | Childhood outcomes:<br>1) Child BMI z-scores at age 2.5<br>2) child BMI z-scores at age 4<br>3) Difference in BMI from age 2.5 to 4<br>4) Child adiposity (skinfolts) at age 2.5<br>5) Child adiposity (skinfolts) at age 4<br>6) Difference in adiposity from age 2.5 to 4 | 1) -0.07 (p=0.56)<br>2) -0.22 (p=0.07)<br>3) 0.11 (p=0.41)<br>4) 0.00 (p=0.97)<br>5) -0.06 (p=0.72)<br>6) 0.03 (p=0.82) |
| Goudet, 2011 (13) | Cohort | Bangladesh | 1998 Bangladesh Flood | 1998 | 220 | Maternal malnutrition among mothers of infants and young children following flood exposure defined as underweight (BMI<18.5 kg/m2) compared to normal (BMI>=18.5) | 12 months | Child outcomes at 12-36 months of age:<br>1) Underweight (weight for age z-score <-2)<br>2) Stunted (height for age z-score <-2)<br>3) Wasted (weight for height z-score <-2) | 1) Adjusted OR = 3.509, 95%CI: 1.022,12.048)<br>2) Adjusted OR: 4.447, 95%CI: 1.044,18.943<br>3) Adjusted OR: 2.097, 95%CI: 0.507, 8.671 |
| Kroska, 2018 (19) | Longitudinal study | USA | Iowa Flood | 2008 | 103 | Levels of maternal stress among those exposed to Iowa floods | 2.5 years | Children outcomes at 2.5 years:<br>BMI | Standardized coefficient: 0.2071 (p=0.0322) |
| Liu, 2016 (23) | Longitudinal study | Canada | Quebec Ice Storm | 1998 | 52-111 at different time points | Levels of maternal stress (objective hardship and subjective stress) among those exposed to Iowa floods | 5.5-15.5 years | Children outcomes at 5.5-15.5 years:<br>1) BMI<br>2) Waist to height ratio | Estimate (95% CI)<br>1) Objective hardship: -0.059 (95% CI: -0.175, 0.058)<br>2) Objective hardship: 0.010 (95% CI: -0.002, 0.021)<br>Subjective stress: 0.044 (95% CI: -0.010, 0.097) |
| Mazumder, 2010<sup>2</sup> (36) | Cohort | USA | 1918 Influenza Pandemic | 1918-1919 | 101,068 | Infants who were born during the Influenza pandemic (third and fourth quarter of 1918, and the first, second and third quarter of 1919) compared to those born in the last quarter of 1919 | ~ 60-82 years | Adult outcomes at 60-82 years:<br>1) Diabetes<br>2) Heart disease | Excess cases of diabetes/heartburn:<br>1) 1918 Q4: 7.7% excess (95% CI: -10.6, 25.9); 1919 Q1: -5.2 (95% CI: -22.9, 12.5); 1919 Q2: 36.7% excess (95% CI: 18.9, 54.4);<br>2) 1918 Q4: 4.6% excess (95% CI: -4.3, 13.5); 1919 Q1: 10.9% excess (95% CI: 2.3, 19.6); 1919 Q2: 6.4 excess (95% CI: -2.2, 15.1); |
| Mryskyla, 2013<sup>2</sup> (37) | Cohort | USA | 1918 Influenza Pandemic | 1918-1919 | 81,571 | Infants who were born during the Influenza pandemic (born during different quarters of 1917, 1918 and 1919) compared to those born in 1920-1924 | ~ 63-95 years | Adult outcomes at 63-95 years:<br>Cardiovascular mortality | 1918 Q1: HR: 1.05 (95% CI: 0.94, 1.17);<br>1918 Q2: HR: 1.02 (95% CI: 0.91, 1.14);<br>1918 Q3 HR: 0.99 (95% CI: 0.89, 1.10);<br>1918 Q4 HR: 0.97 (95% CI: 0.87, 1.09);<br>1919 Q1 HR 1.07 (95% CI: 0.96, 1.19);<br>1919 Q2: HR:1.06 (95% CI: 0.95, 1.19); |
| Oni, 2015 (25) | Cross-sectional | USA | Hurricane Katrina | 2005 | 146 | Women who were pregnant during Hurricane Katrina or became pregnant immediately after hurricane compared to those who were not exposed to the hurricane; Women who experienced prenatal stress caused by Hurricane Katrina compared to those who did not experience stress | 9 months | Childhood outcomes at 9 months:<br>1) Pregnancy induced hypertension<br>2) Gestational diabetes | 1) Hurricane exposure: adjusted OR: 1.22 (95% CI: 0.81, 1.84); perceived stress: adjusted OR: 1.16 (95% CI: 1.05, 1.30);<br>2) Hurricane exposure: adjusted OR: 1.04 (95% CI: 0.69,1.57); perceived stress: adjusted OR: 1.13 (95% CI: 1.02, 1.25) |
| Sotomayer, 2013 (40) | Cohort<br>Natural experiment | Puerto Rico | Hurricane San Felipe and San Ciprian | 1928 and 1932 | 11990 | Those born during 1929 and 1933 were defined as exposed to the hurricanes compared to individuals born outside of these years between 1920 and 1940 | Not reported (average age = 70) | Outcomes at ~70 years of age:<br>1) Diabetes<br>2) Hypertension<br>3) High cholesterol<br>4) CVD<br>5) AMI<br>6) Coronary/angina<br>7) Stroke | Parametric estimates (SE)<br>1) San Felipe: 6.77 (0.98); San Ciprian: 4.22 (0.98)<br>2) San Felipe: 6.08 (1.07); San Ciprian: 5.63 (1.07)<br>3) San Felipe: 10.06 (1.21); San Ciprian: 4.67 (1.21)<br>4) San Felipe: -1.84 (2.02); San Ciprian: 2.23 (2.02)<br>5) San Felipe: 1.08 (1.25); San Ciprian: 2.95 (1.25)<br>6) San Felipe: 0.13 (2.45); San Ciprian: 0.60 (2.45)<br>7) San Felipe: 0 (0.83); San Ciprian: 0.96 (0.83) |
| Xiao, 2019 (26) | Time series/Quasi-experimental | USA | Hurricane Sandy | 2012 | Not reported | Exposure to Hurricane Sandy lasting impacts defined as the following 12 months after Sandy (November 2012-October 2013) compared to the November to October in other years during November 2005 to October 2014 among women who were pregnant | 12 months | Outcomes in adults:<br>1) Emergency department visits for gestational hypertension<br>2) Emergency department visits for diabetes or abnormal glucose | 1) Increased at 7 months: 7.3% (95% CI: 1.0%, 13.9%)<br>2) Increased at 8 months:26.3% (95% CI: 3.9%, 53.6%)<br>*results for 12 month reported graphically |

1. Only presenting results for early and late gestation; results for mid gestation are not included in summary table but can be found in studies 2. Not all results presented for different exposure groups
Abbreviations: AMI: Acute myocardial infarction; BMI: Body mass index; BP: Blood pressure; CI: Confidence Interval; CHD: Coronary heart disease; Chol: Cholesterol; CVD: Cardiovascular disease; HDL: High-density lipoproteins; HR: Hazard ratio; LDL: Low-density lipoproteins OR: Odds Ratio; PMNS: Prenatal Maternal Stress; Q1-Q4: quarter; SE: Standard error; Trig; Triglycerides.

**Table 4.** Description of studies investigating the association between exposure to a disaster during adulthood and cardiometabolic outcomes across the life-course, by disaster type (n=34)

| Study | Study design | Country | Name of disaster | Year | Sample size | Primary exposure and comparator | Average follow-up | Outcomes | Primary results |
| --- | --- | --- | --- | --- | --- | --- | --- | --- | --- |
| **Human-made disaster** |  |  |  |  |  |  |  |  |  |
| Brackbill, 2006 (68) | Cohort | USA | World Trade Center attacks (9/11) | 2001 | 8,418 | Adult survivors of 9/11 present at time of first airplane impact in a structure that was damaged compared to those that collapsed; time of evacuation before compared to after damage | 1 year | 1) Hypertension<br>2) Coronary heart disease<br>3) Angina<br>4) Heart attack<br>5) Diabetes<br>6) Stroke | 1) Building type: adjusted OR: 1.2 (p<0.05); time of evacuation: adjusted OR: 0.9 (0.6, 1.3)<br>2) Building type: adjusted OR: 0.8 (0.4, 1.6); time of evacuation: adjusted OR: 0.5 (0.1, 2.2)<br>3) Building type: adjusted OR: 0.8 (0.4, 1.6); time of evacuation: adjusted OR: 0.7 (0.2, 3.1)<br>4) Building type: adjusted OR: 2.1 (0.9, 4.9);<br>5) Time of evacuation: adjusted OR: 0.7 (0.3, 1.7)<br>6) Building type: adjusted OR: 1.5 (0.6, 4.0) |
| Dirkzwager, 2007 (51) | Cohort | Netherlands | Fireworks depot explosion | 2000 | 896 | PTSD among those exposed to the fireworks disaster 19 months following the disaster compared to those with no PTSD exposed to the fireworks explosion | 18 months | 1) Cardiovascular<br>2) Vascular problems | 1) Physical health problems OR: 1.23; 95% CI: 0.78, 1.94; New health problems (not present pre disaster): 1.11; 0.65, 1.89<br>2) Physical health problems OR: 2.12 95% CI: 1.23, 3.68; 1.92; New health problems (not present pre disaster) OR: 1.92, 95% CI: 1.04, 3.55. |
| Dorn, 2007 (41) | Cohort | Netherlands | Volendam Pub Fire | 2001 | 2255 | Parents of children with burns from fire parents of children without burns, bereaved parents compared to community controls who were not trapped in fire | 4 years | Incidence of hypertension | Bereaved parents: OR: 2.42, 95%CI: 0.90, 6.55); parents of victims with burns: OR: 1.43, 95%CI: 0.97, 2.11; parents of victims without burns: OR: 1.44, 95%CI: 0.92, 2.26) |
| Gerin, 2005 (69) | Pre/post design/quasi experimental | USA | World Trade Center attacks (9/11) | 2001 | 528 | Adults 2 months before 9/11 compared to 2 months after 9/11 across 4 cities (Chicago, Washington DC, New York and Mississippi) | 4 months | Systolic blood pressure | Difference (SE)<br>New York: 1.58 (0.82) p<0.05<br>Chicago: 2.15 (0.32) p<0.001<br>Mississippi: 2.92 (0.67) p<0.001<br>Washington DC: 8.67 (1.16) p<0.001 |
| Huizink, 2006 (60) | Cohort | Netherlands | Amsterdam Air Disaster | 1992 | 1996 | Police officers and firefighters who performed at least one disaster-related task compared to professional colleagues who did not perform any disaster-related tasks | 8.5 years | Cardiovascular complaints | Adulthood outcomes<br>Police officers: OR: 1.76 (95% CI: 1.35, 2.29)<br>Firefighters: OR: 3.3 (95% CI: 1.70, 6.41) |
| Jordan, 2011A (55) | Prospective cohort | USA | World Trade Center attacks (9/11) | 2001 | 39,324 | 9/11-related PTSD compared to no PTSD | 2.9 years | Heart disease | Women adjusted OR: 1.68 (95% CI: 1.33, 2.12)<br>Men adjusted OR: 1.62 (95% CI: 1.34, 1.96) |
| Jordan, 2011B (61) | Prospective cohort study | USA | World Trade Center attacks (9/11) | 2001 | 39324 | Low, intermediate and high exposure to 9/11 | 2.9 years | Heart disease mortality | Intermediate exposure: HR: 1.21 (95% CI: 0.80, 1.83)<br>High exposure: HR: 2.06 (95% CI: 1.10, 3.86). |
| Jordan, 2013<sup>1</sup> (47) | Cohort | USA | World Trade Center attacks (9/11) | 2001 | 46,346 | Low, intermediate and high exposure to 9/11 | 7 years | CVD hospitalizations | Rescue/recovery workers: women: high: adjusted HR: 3.29 (95% CI: 0.85, 12.69); men: high: 1.82 (95% CI: 1.06, 3.13)<br>Non-rescue/recovery workers: women: high: adjusted HR: 0.88 (95% CI: 0.54, 1.43); men: high: adjusted HR: 0.94 (95% CI: 0.60, 1.47) |
| Kong, 2019 (48) | Pre/post design/quasi experimental | South Korea | Sewol Ferry Disaster | 2014 | 73,632 | Exposure to the Sewol Ferry Disaster in one-week periods from May 21 through June 17, 2014 compared to the reference period (March 2015-April 2015) | 8 weeks | Adulthood outcomes<br>1) Acute MI<br>2) Angina | 1) 8 weeks after Sewol: IRR: 0.91 (95% CI: 0.81, 1.02)<br>2) 8 weeks after Seowl: IRR: 0.93 (95% CI: 0.85, 1.01) |
| Lin, 2010 (66) | Pre/post design/quasi experimental | USA | World Trade Center attacks (9/11) | 2001 | Not reported | Areas affected by 9/11 compared to areas not affected by 9/11 | 10 years | Adulthood outcomes for Cardiovascular disease hospitalizations | Prevalence ratio (95% CI):<br>08/14–09/10: 0.51 (95% CI: 0.26, 1.00)<br>09/11–09/17: 0.56 (95% CI: 0.28-1.11);<br>09/18-09/24: 0.77 (95% CI: 0.44, 1.32);<br>09/25-10/01: 0.49 (95% CI: 0.24, 1.00);<br>10/02-10/08: 0.98 (95% CI: 0.53, 1.87);<br>10/09-10/15: 1.09 (95% CI: 0.60, 1.98);<br>10/16–10/22: 0.50 (95% CI: 0.26-0.95);<br>10/23–10/29: : 0.45 (95% CI: 0.20-0.98);<br>10/30–11/05: 0.48 (95% CI: 0.23-0.97) |
| Yu, 2018 (67) | Cohort Study | USA | World Trade Center attacks (9/11) | 2001 | 42,527 | 9/11-related PTSD compared to no PTSD | 13 years | Stroke | Adjusted HR: 1.69 (95% CI: 1.42, 2.02) |
| **Natural disaster** |  |  |  |  |  |  |  |  |  |
| An, 2015 (65) | Cross-sectional | USA | Hurricane Ike | 2008 | 19 | Psychological strains among Hurricane Ike survivors | 3 months | 1) Blood glucose<br>2) Obesity | Mean (high vs low) and standard deviation:<br>PTSD symptom:22.44 (4.93) vs.12.86 (10.48);<br>p=0.014; perceived stress:23.00 (5.03) vs. 28.11 (5.07) p=0.048<br>2) 28.43 kg/m<sup>2</sup> (3.92) vs. 20.83kg/m<sup>2</sup> (3.92) p=0.018 |
| Baum, 2019 (12) | Cohort study | USA | Hurricane Sandy | 2012 | 81 544 | Veterans who used Manhattan VA Medical Center before Hurricane Sandy and experienced decreased access to health care services compared to veterans who used the VA Bronx, Brooklyn or West Haven medical centers | 2 years | 1) Uncontrolled hypertension<br>2) Systolic BP (mmHg)<br>3) Diastolic BP (mmHg)<br>4) Uncontrolled diabetes<br>5) Uncontrolled cholesterol<br>6) Weight | % differential change (95% CI):<br>1) 6 months: 19.3 (4.5, 8.7); 12 months: 4.5 (3.1, 5.9); 18 months: 5.0 (3.5, 6.5); 24 months: 2.1 (0.5, 3.6)<br>2) 6 months: 3.8 (3.1, 4.5); 12 months: 2.3 (1.7, 2.9); 18 months: 3.1 (2.5, 3.7); 1.5 (0.9, 2.1)<br>3) 6 months: 2.7 (2.3, 3.1); 12 months: 2.2 (1.9, 2.6); 18 months 2.9 (2.5, 3.3), 24 months: 2.0 (1.7, 2.4)<br>4) 6 months: 1.9 (-0.1, 4.0); 12 months: 1.7 (-0.3, 3.6); 18 months: 0.8 (-1.2, 2.8); 24 months: -0.2 (-2.2, 1.8)<br>5) 6 months: 1.3 (-0.1, 2.6); 12 months: 0.6 (-0.6, 1.8); 18 months: -0.7 (-2.0, 0.6); 24 months: -0.2 (-1.4, 1.0)<br>6) 6 months: -0.1 (-0.5, 0.2); 12 months: 0.2 (-0.2, 0.5); 18 months: -0.2 (-0.5, 0.2); 24 months: 0.5 (0.1, 0.9) |
| Becquart, 2018 (46) | Time series/quasi experimental | USA | Hurricane Katrina | 2005 | 383 552 | Exposure to hurricane before, during and after among older adults in Louisiana in the affected counties | 1 year | Adult outcomes<br>Hospitalizations due to CVD | Mean (SD)<br>Orleans: T1: 7.25 (2.44); T2: 3.91 (1.45)*; T3: 18.47 (17.3)*; T4:13.76 (6.51)*; T5: 9.54 (2.78); T6: 4.69 (2.08)<br>Jefferson: T1: 5.90 (1.90); T2: 5.01(1.52); T3: 8.118 (3.70)*; T4: 7.25 (2.15)*; T5: 5.26 (1.53); T6: 4.65 (1.57)*<br>East BR: T1: 8.69 (2.74); T2: 9.11 (2.69); T3: 6.52 (2.58); T4: 6.55 (1.70)*; T5: 6.69 (2.42)*; T6: 7.39 |
|  |  |  |  |  |  |  |  |  | (2.37)*<br>* p<0.05 |
| Bich, 2011 (59) | Cross-sectional | Vietnam | Historic flood in 2008 | 2008 | 781 | Individuals who resided in households affected by flood in Hanoi in 2005 compared to non-affected households | 1 month | Worsening hypertension after rain/flood | Rural: non flooded 33.3%; flooded: 51.2%; Urban: non flooded 20.3% flooded: 42.9%*<br>* p<0.05 |
| Fonseca, 2009 (57) | Cohort | USA | Hurricane Katrina | 2005 | 1795 | Adults with diabetes who were in the databases from 3 health care systems 6 months before the hurricane (Feb 28, 2005-Aug 27, 2005) compared to 6-16 months after the hurricane (March 1, 2006-December 31, 2006) | 22 months | 1) Glycemic control/A1C<br>2) Systolic BP (mmHg?)<br>3) Diastolic BP (mmHg)<br>4) HDL<br>5) LDL<br>6) Triglycerides | Difference in mean (SD)<br>1) 0.1 (1.6) (p<0.01)<br>2) 10.5 (20.4) (p<0.01)<br>3) 3.9 (13.1) (p<0.01)<br>4) 6.0 (35.5) (p<0.01)<br>5) -2.4 (9.2) (p<0.01)<br>6.) -2.1 (137.5) (p=0.60) |
| Gautam, 2009 (53) | retrospective cohort | USA | Hurricane Katrina | 2005 | 396 | Exposure to Hurricane Katrina compared to period before hurricane | 4 years | Incidence of AMI admission | Pre-Katrina group: 150 admissions for AMI (0.71%)<br>Post-Katrina group: 246 admission for AMI (2.18%)<br>p<0.0001 |
| Hendrickson, 2996 (62) | Pre/post design/quasi experimental | USA | Hurricane Iniki | 1992 | Not reported | Mortality data for residents of Kauai for 5-year period 1987-1991 prior disaster compared to the year immediately following the hurricane (Oct 1 1992-Sept 30, 1993) | 6 years total | Mortality by:<br>1) Heart disease<br>2) Stroke<br>3) Diabetes mellitus | 1) RR: 0.96 (95% CI: 0.79-1.17)<br>2) RR: 1.20 (95% CI: 0.81-1.78)<br>3) RR: 2.61 (95% CI: 1.44-4.74) |
| Husarewycz, 2014 (70) | Cross-sectional | USA | Natural disaster/terrorism | Lifetime disaster experience | 34,653 | Number of times directly experienced natural disaster/terrorism compared to no experiences | 1 year | 1) Cardiovascular disease<br>2) Hypertension/arteriosclerosis<br>3) Diabetes<br>4) Obesity | 1) OR: 1.28 (95% CI: 1.10, 1.49)<br>2) OR: 1.08 (95% CI: 0.95, 1.24)<br>3) OR: 1.10 (95% CI: 0.94, 1.29)<br>4) OR: 1.01 (95% CI: 0.90, 1.14) |
| Jiao, 2012 (52) | retrospective cohort observational study | USA | Hurricane Katrina | 2005 | Not reported | 2 years prior to the hurricane (August 29, 2003 - August 28, 2005) compared to the 3-year period post-Hurricane Katrina (February 14, 2006 - February 13, 2009) | 5 years | Incidence of AMI | Pre-Katrina: 0.7% compared to post-Katrina: 2% (p<0.001) |
| Joseph, 2014 (49) | Cohort/longitudinal | USA | Hurricane Katrina | 2005 | 215 | African Americans who experienced acute unemployment due to Hurricane compared to those who remained employed | 4 years | Cardiometabolic event | Adjusted OR = 5.65, p < .05 |
| Karatzias, 2015 (42) | Cross-sectional | Hong Kong | Natural disaster | Not specified | 1147 | Experience of natural disaster across life-course compared to less or no experiences | Survey done from August to December 2012 | 1) Hypertension<br>2) Heart disease<br>3) Diabetes | Chi square (p-value)<br>1) X2: 3.3 p=0.047<br>2) X2: 3.6 p=0.056<br>3) X2: 2.5 p=0.088 |
| Kim, 2017 (56) | Pre/post design/quasi experimental | USA | Hurricane Sandy | 2012 | Not reported | The month of Hurricane Sandy (October 28, 2012-November 27, 2012) compared to the same month in 2009-2011; Sandy quarter (October 28, 2012-January 27, 2013) compared to the same period in 2009-2011 among elderly people | Sandy month: Oct 28, 2012-Nov 27, 2012<br>Sandy quarter: Oct 28, 2012-Jan 27, 2013 | Cardiovascular disease-related death | Sandy quarter: adjusted RR: 1.06; 95% CI: 1.02, 1.10<br>Sandy month: adjusted RR: 1.10; 95% CI: 1.02, 1.18 |
| Koroma, 2019 (64) | Cross-sectional | Sierra-Leone | Ebola | 2014-2015 | 10011 | District facilities for six-month periods before Ebola (June-December 2012), during Ebola (June-December 2014) and post-Ebola (June-December 2015) | June-December 2012,2013,2014 | 1) Cardiovascular disease<br>2) Hypertension<br>3) Diabetes | 1) Pre-Ebola: 355, Ebola: 300, Post-Ebola: 196<br>2) Pre-Ebola: 282, Ebola: 230, Post-Ebola: 457<br>3) Pre-Ebola: 3716, Ebola: 1851, Post-Ebola: 2463 |
| Lawrence, 2019 (43) | Prospective Cohort Study | USA | Superstorm Sandy | 2012 | 651858 | Residing in counties affected by Superstorm Sandy compared to non-affected counties; Superstorm period compared to reference periods (short-term and long-term (4 and 12 months)) | 1 year | Emergency department visits, outpatient visits, and hospital admissions for cardiovascular disease | 4 months: Superstorm sandy period: RR: 2.10 (95% CI: 2.10, 2.10); Affected counties RR: 2.62 (95% CI: 2.62, 2.63)<br>12 months: Superstorm sandy period: RR: 2.01 (95% CI: 2.00, 2.01); Affected counties RR: 2.64 (95% CI: 2.64, 2.65) |
| McKinney, 2011 (50) | Time-series/quasi experimental | USA | Hurricane Charley, Frances, Ivan and Jeanne and Tropical Storm Bonnie | 2004 | Not reported | Counties in 2004 directly impacted by the hurricanes, ordered evacuated regardless of the level of damage that occurred and adjacent to the impact zone where direct deaths were reported compared to compared to the same areas in 2001-2006 | 5 years | Heart-related mortality | Results shown graphically<br>Significantly elevated heart-related deaths |
| Moscona, 2019 (73) | Retrospective cohort study | USA | Hurricane Katrina | 2005 | 2-year pre-Katrina - 21,079<br>10-year post Katrina - 84,751 | Individuals who lived in New Orleans who went to the Tulane University Health Sciences Center compared to the two months prior to the Hurricane | 12 years | 1) Hospital admission for Incidence of AMI<br>2) Changes in CAD<br>3) Changes in diabetes mellitus<br>4) Changes in hypertension<br>5) Changes in hyperlipidemia | Pre-Katrina versus Post-Katrina<br>1) 0.7% vs 2.8% (p<0.001)<br>2) 36.4% vs. 47.9%, (p= 0.01)<br>3) 31.3% vs. 39.9% (p= 0.04)<br>4) 71.1% vs 80.6%, (p=0.12)<br>5) 45.4% vs. 59.3% (p = 0.005) |
| Nagayoshi, 2015 (54) | Pre/post design/quasi experimental | Japan | July 12, 2012 heavy rain and mudslides "mountain tsunamis" | 2012 | 583 | Individuals who were admitted at Aso Central Hospital from July 12 to August 31, 2012 compared to the 3-year period before flooding | 3 years | 1) Hospital admission for cardiovascular outcomes<br>2) Cardiovascular events | 1) 4.5/month before compared to 16.8/month after; p < 0.01<br>2) 5.1/month before compared to 16.8/month after; p < 0.01. |
| Ng, 2011 (71) | Cohort | United Kingdom | Flood | June, 2007 | 1,743 | Diabetics affected by floods compared to diabetics not affected by floods | 2 years | glycemic control/HbA1c levels | Mean HbA1c before 7.6% (7.5–7.7) vs. After 7.9% (7.7–8.0); p = 0.002 |
| Peters, 2013 (63) | Retrospective Cohort | USA | Hurricane Katrina | 2005 | 698 | Admission to Tulane University Health Sciences Centre in the 3-year period post-Katrina compared to the 6-year period pre-Katrina | 9 years | Chronobiology of AMI onset | Pre-Katrina: 45% vs post-Katrina: 30.9%, p=0.002 |
| Rey, 2007 (72) | Longitudinal | France | 6 Heat Waves | 1971-2003 | Not reported | Time of heat wave compared to the expected mortality during the 3 years prior to the heat wave | N/A | Excess cardiovascular disease death | 41% in 1975 to 23% in 2003 |
| Sliva-Palacios, 2015 (58) | Pre/post design/quasi experimental | USA | Oklahoma Tornado | 2013 | 22,607 | Victims of the Oklahoma Tornado Outbreaks compared to the same people pre-tornado and same period one year prior | 6 Months | Hospital admissions for CVE | One year prior: PR = 1.05 95% CI: 0.91 to 1.21, p = 0.50; 3-month pre-tornado: PR = 0.96, 95% CI: 0.83 to 1.21, p = 0.63 |
| Thethi, 2010 (74) | Cohort | USA | Hurricane Katrina | 2005 | 1523 | Individuals exposed to Hurricane Katrina compared to 6-16 months pre-Hurricane Katrina (February 28, 2005-August 27, 2005) | 6 months before Katrina and 6-16 months after Katrina and follow up 1 year after the first post-Katrina visit | 1) LDL<br>2) HDL<br>3) Triglycerides<br>4) Cholesterol<br>5) Diastolic blood pressure<br>6) Systolic blood pressure | Pre-Katrina vs Post-Katrina mean:<br>1) 101.34 vs 107.44<br>2) 43.53 vs 41.08<br>3) 160.8 vs 158.65<br>4) 181.9vs 181.39<br>5) 70.99 vs 74.88<br>6) 130.73 vs 141.27 |
| Vanasse, 2016<br>(44) | Population-based retrospective cohort study with a time series design | Canada | Flood of Saint-Jean-sur-Richelieu | 2011 | 111,317 | Exposure to flood in spring 2011 and exposure to flooded area (Area 1) compared to same period in Spring 2010 and 2012 and non-flooded areas in the same town (Areas 2, 3 and 4) | 4 months | Acute cardiovascular event | Spring 2010: aOR 1.25 (95%CI: 0.81, 1.92); spring 2012 aOR: 1.27 (95% CI: 0.82, 1.92); Non-flooded areas 2: aOR: 1.11 (95%CI: 0.79, 1.59), Non-flooded area 3: aOR: 0.94 (95% CI: 0.68, 1.32); Non-flooded area 4: aOR 1.08 (95%CI: 0.78, 1.47) |

1. Only results for extreme outcomes are reported in table, remaining results can be found in the study.
Abbreviations: aOR: Adjusted odds ratio; AMI: Acute myocardial infarction BMI: Body mass index; BP: Blood pressure; CI: Confidence Interval; CAD: Coronary artery disease; CHD: Coronary heart disease; CVD: Cardiovascular disease; CVE: Cardiovascular events; HbA1c ; Hemoglobin A1C; HDL: High-density lipoproteins; HR: Hazard ratio; LDL: Low-density lipoproteins OR: Odds Ratio; PR: Prevalence Ratio; PTSD: Post-traumatic stress disorder; RR: Relative risk; SD: Standard deviation; SE: Standard Error; Trig: Triglycerides.; T1-T6; Time.

### Studies on perinatal and childhood exposure to disaster and subsequent cardiometabolic outcomes

Of the 24 studies that evaluated perinatal and childhood exposure to disaster, 12 studies evaluated human-made disasters (27–35,38,39,45) and the remaining 12 evaluated natural disasters (13,19–26,36,37,40) of which two were pandemics (36,37). Most studies (n=15) assessed the disaster as the main exposure of interest (22,26–31,33–37,37–40). The remaining studies evaluated stress (e.g., maternal stress, disaster-related PTSD, subjective stress, objective hardship) (19–23,25,45), maternal weight and maternal nutrition status (13,79), cognitive appraisal (24), and coping strategies (25) that were the result of the disaster as the exposure variable. The age when cardiometabolic outcomes were assessed varied across studies, with 7 studies evaluating outcomes during childhood and adolescence (≤18 years of age) (13,19–24), 4 studies during young to mid adulthood (>18-40 years of age) (25,34,80,83), and 10 during later adulthood (≥50 years of age) (28,30–33,36–40). Ekamper et al. (2015), evaluated outcomes throughout the life-course, with age at outcome ranging from 18-63 years (29). Two studies (26,45) did not specifically identify the age at which outcomes were assessed. Detailed characteristics and findings of all studies that assessed perinatal and childhood exposures to disasters can be found in Table 3.

Studies that assessed human-made disasters found an increased association or increased change in obesity, metabolic syndrome, CVD related-outcomes and diabetes, however not all were statistically significant (OR range 1.15-3.11; hazard ratio (HR) range: 1.26-1.91) (27,29–35,38,45). Some studies also reported no association or a decreased association following exposure to human-made disasters and cardiometabolic outcomes (OR range: 0.94-1.02) (29,34). Among the studies that evaluated natural disasters, maternal stress, including objective hardship and objective prenatal maternal stress, were found to increase the odds of obesity using different adiposity measures during childhood and adolescence (19–21). However, weak correlations were found between subjective distress/stress and body mass index (BMI) (r=0.39; p=0.03), zBMI (r=0.40; p=0.02), and body fat percentage (r=0.33; p=0.09) in offspring at age 13 (22). The prevalence of heart disease, ischemic heart disease, emergency department visits for gestational hypertension, myocardial infarction, diabetes diagnoses and diabetes-related emergency department visits increased after exposure to Hurricanes Sandy, San Felipe and Ciprian, and the 1918 Influenza Pandemic (26,36,40). In addition, exposure to hurricanes and stress-related to hurricanes was noted to increase pregnancy induced hypertension and gestational diabetes (odds ratio (OR) range: 1.13-1.22) (25).

### Studies on adult exposure to disaster and subsequent cardiometabolic outcomes

Thirty-four studies investigated the effects of exposure to disasters during adulthood on cardiometabolic outcomes. The length of follow-up ranged from 1 month to 13 years. There were 23 studies that examined natural disasters (12,42–44,46,49,50,52–54,56–59,62,63,65,71–74), and 11 studies that examined human-made disasters (41,47,48,51,55,60,61,66–70). Of these studies, only one evaluated the impact of an infectious disease epidemic (64). Most studies (12,41–44,46,48,50,52–54,56–64,66,69–74) considered the disaster as the main exposure of interest (n=27). The remaining 7 studies assessed disaster-related stress (51,55,65,67), including post-traumatic stress disorder (PTSD) and psychological strain, unemployment rates as a result of the disaster (49) and exposure to damaged or collapsed buildings during the World Trade Center disaster (68). Detailed characteristics and findings of all studies that assessed adult exposures to disasters are included in Table 4.

The studies that assessed exposure to human-made disasters during adulthood reported mixed results in terms of associations with outcomes and statistical significance. Most studies reported an increased association with outcomes, including heart disease-related mortality, CVD hospitalization, complaints of CVD, vascular problems, stroke, and incidence of hypertension (OR range: 1.21-3.30; HR range: 1.63-1.82) (41,43,47,51,55,60,61,67,68,70). No association or a decreased association with CVD hospitalizations was noted following exposure to the World Trade Centre attacks (47,66). In addition, time of evaluation from damaged buildings resulting from the World Trade Centre also had a decreased association with cardiometabolic outcomes (OR range: 0.7-0.9) (68). Among studies that evaluated the impact of exposure to natural disasters on subsequent cardiometabolic outcomes, nearly all studies reported an increased association (OR range: 1.08-5.65; relative risk (RR) range: 2.01-2.62; HR range: 1.72-3.29) (43,44,49,56,62,70). However, five studies reported no association or a decreased association after exposure to hurricanes among specific groups or at different timepoints following a disaster (47,56,58,62,70). Several studies evaluated the mean difference in outcomes including cardiovascular, diabetes and obesity-related outcomes, however, these results were very inconsistent, with some studies noting an increase, some a decrease and other reporting no change in outcomes following a disaster (12,46,52–54,57,59,63,65,73,74).

### Mediation and modification of cardiometabolic outcomes

Across all studies, few evaluated effect modification or subgroups of a population that may be at a greater risk of negative health outcomes following disasters. Some studies stratified by sex (28,32,34,39,43,48,55,62), gestational timing of exposure (20,27,29–31,33,36), year of birth or age at outcome (40,62,67), urban or rural area (35), race (43,46) and socioeconomic status (65), however, results varied greatly due to the differences in exposure period, disaster type, cardiometabolic outcome and age at outcome. One study explored the possible mediators between cognitive appraisal following the Quebec Ice Storm and obesity. It was noted that negative cognitive appraisal was found to predict obesity via DNA methylation of diabetes-related genes (24). No studies evaluated or discussed possible interventions to mitigate risk of cardiometabolic disease following a disaster.

### Critical appraisal

The critical appraisal assessment for all study designs are found in Tables A2-A4. Among the cohort studies, most studies met all criteria included in the checklist indicating high study quality. For instance, almost all cohort studies had comparable populations that were recruited in a similar way and exposures that were assessed in the same way across populations. However, across almost all cohort studies, information on follow-up or strategies to address incomplete follow-up were unclear or not addressed. The critical appraisal results for cross-sectional studies were inconsistent with a small number of studies meeting only some checklist requirements. For quasi-experimental studies, the checklist requirement for within person comparisons were not applicable for all studies, however, all studies clearly defined the cause and effect within the study.

## DISCUSSION

Exposure to both natural and human-made disasters, including famine, war, terrorism, natural disasters, and infectious disease epidemics, during both the perinatal/childhood and adult periods were associated with increased cardiometabolic outcomes including obesity, hypertension, myocardial infarction, diabetes, and cardiac mortality in most studies. These findings emphasize that the burden of disasters extends beyond the known direct harm they cause, and attention is needed on the detrimental indirect long-term effects on cardiometabolic health and chronic disease. Meta-analysis was not possible due to the very heterogeneous study designs, measures of effect, outcomes, and timing of exposures. The included studies also reported varied follow-up periods and length of retrospective assessment. From an epidemiological standpoint, the difference in the follow-up periods, timing of exposure, year of disaster and geographic location may help to explain the variation in the reported results. It is important for future studies to explore these differences and how they may influence cardiometabolic outcomes.

Studies also reported insufficient data on subgroups that were at increased risk of worse cardiometabolic health outcomes and interventions that were implemented to mitigate risk of cardiometabolic outcomes. There were also few results that investigated sex and gender differences, or that applied an equity lens. It has been noted that those of different levels of socioeconomic status experience differential cardiometabolic outcomes (84,85). This signifies the importance of exploring associations between exposure to disasters and cardiometabolic outcomes stratified by these factors. Understanding how these associations differ will also help to identify groups of people who will experience worse outcomes following a disaster.

The potential mechanisms discussed whereby exposure to disaster could increase cardiometabolic outcomes include maternal objective hardship, subjective stress and various coping mechanisms, psychological strains including PTSD, anxiety and fear, and reduced health care service access. One study that explored mediators in the association between stress and obesity measures identified the role of DNA methylation in this association (24). The findings from this study are important to better understand a possible pathway of obesity development following exposure to a disaster. More studies are needed that evaluate other mechanisms, such as disruptions to healthcare or medication access, and changes in dietary intake or physical activity. Understanding the mechanistic pathway following exposure to disaster will allow for targeted public health strategies, contributing to a mitigation of risk.

This review comprehensively evaluated of the impact of a wide range of disaster exposures on various cardiometabolic outcomes across the life-course. The search strategy was developed in consultation with health science librarians at McMaster University to ensure the most comprehensive search was developed and relevant literature was identified. Very few studies have evaluated the long-term impacts of pandemics and epidemics on cardiometabolic outcomes, identifying a current gap in the literature. The timely findings of this synthesis are a strength of this review, given the current COVID-19 pandemic, which is affecting millions of people worldwide. While only a small proportion of the identified studies focused on pandemics and epidemics, the findings may serve to guide our understanding of expected outcomes and to develop future research to study the effects of COVID-19 on cardiometabolic outcomes. The results from this study can also be used to better understand the trade-off between the implementation of public health measures, such as physical distancing to reduce transmission of a virus, and the implications with access to healthcare, as the review found access to healthcare was limited following a disaster (12). The findings from this review can help to inform future decisions by policy makers regarding the implementation of various public health measures.

The overall findings of this review indicate exposure to any type of disaster may increase the risk of cardiometabolic outcomes including obesity, diabetes and CVD. Further, our results suggest that increased risk is observed for disaster exposure at any period over the life-course from the perinatal child and adult periods. However, more studies are needed to understand the mechanisms which may explain these associations and to identify subgroups of the population at highest risk of long-term cardiometabolic outcomes post-disaster. Findings from this review may inform research that should be conducted in the context of the current COVID-19 pandemic and may inform public health mitigation measures that should be considered post-pandemic to prevent long term cardiometabolic outcomes in the population.

## Supporting information

PRISMA Checklist

## Data Availability

All data is available in the manuscript or supplementary material.

## Author contributions

Conceptualization: V.D., S.E.N.S., E.A., E.A., L.N.A.; Screening: V.D., J.L., M.S.A., Y.Y.M., A.T.A., E.S., S.I., J.D.M., R.R.; Data extraction: V.D., J.L., M.S.A., Y.Y.M., A.T.A., R.R.; Critical appraisal: V.D., J.L., M.S.A., Y.Y.M., A.T.A., E.S., S.I.; Writing – Original draft: V.D., J.L., M.S.A., Writing – review and editing: V.D., J.L., M.S.A., Y.Y.M., A.T.A., E.S., S.I., J.D.M., R.R. S.E.N.S., E.A., E.A., L.N.A.

