## Supplemental Tables 1-4 for "Impact of disasters, including pandemics, on cardiometabolic outcomes across the life-course: A systematic review"

### APPENDIX

**Figure A1.** Search strategy for Medline

| | |
|---|---|
| 1 "social isolation.mp." or Social isolation/ | 1479 |
| 2 quaratine.mp. or exp Quarantine/ | 5589 |
| 3 Pandemics/ | 7247 |
| 4 Epidemics/ | 10146 |
| 5 Coronavirus or coronavirus.mp. | 19310 |
| 6 humanitarian crises.mp. | 232 |
| 7 exp mass casualty incidents/ or exp natural disasters/ | 18883 |
| 8 public health emergency.mp. | 1636 |
| 9 cardiovasucalr.mp. or exp Cardiovascular Diseases/ | 2605399 |
| 10 hypertension.mp. or exp Hypertension/ | 484321 |
| 11 exp Obesity/ or obesity.mp. | 321847 |
| 12 body mass index.mp. or exp Body Mass Index/ | 236149 |
| 13 stroke.mp, or exp Stroke/ | 310109 |
| 14 myocardial infarction.mp. or exp Myocardial Infarction/ | 247417 |
| 15 angina.mp. or exp Angina Pectoris/ | 698186 |
| 16 diabetes.mp. or Diabetes Mellitus/ | 646582 |
| 17 9 or 10 or 11 or 12 or 13 or 14 or 15 or 16 | 3552066 |
| 18 Disasters/ or disaster.mp. | 39131 |
| 19 1 or 2 or 3 or 4 or 5 or 6 or 7 or 8 or 18 | 10917 |
| 20 17 and 19 | 3688 |
| 21 limit 20 to humans | 2998 |

**Table A2.** Critical appraisal using Joanna Briggs Institute (JBI) checklist for cohort studies by exposure period (n=41)

| Author, Year | Were the two groups similar and recruited from the same population? | Were the exposures measured similarly to assign people to both exposed and unexposed groups? | Was the exposure measured in a valid and reliable way? | Were confounding factors identified? | Were strategies to deal with confounding factors stated? | Were the groups/participants free of the outcome at the start of the study (or at the moment of exposure)? | Were the outcomes measured in a valid and reliable way? | Was the follow up time reported and sufficient to be long enough for outcomes to occur? | Was follow up complete, and if not, were the reasons to loss to follow up described and explored? | Were strategies to address incomplete follow up utilized? | Was appropriate statistical analysis used? |
|---|---|---|---|---|---|---|---|---|---|---|---|
| **Perinatal** | | | | | | | | | | | |
| Dancause, 2012 | Yes | Yes | Yes | Yes | Yes | Yes | Yes | Yes | Yes | Yes | Yes |
| Dancause, 2013 | Yes | Yes | Yes | Yes | Yes | Yes | Yes | Yes | No | No | Yes |
| Dancause, 2015 | No | Yes | Yes | Yes | Yes | Yes | Yes | Yes | No | Unclear | Yes |
| de Rooij, 2007 | Yes | Yes | Yes | Yes | Yes | Yes | Yes | Yes | Yes | Yes | Yes |
| Dorn, 2007 | Yes | Yes | Yes | Yes | Yes | Yes | Yes | Yes | Yes | Unclear | Yes |
| Ekamper, 2015 | Yes | Yes | Yes | Yes | Yes | Yes | Yes | Yes | Yes | Yes | Yes |
| Goudet, 2011 | Yes | Yes | Yes | Yes | Yes | Yes | Yes | Yes | Unclear | Unclear | Yes |
| Huang, 2010 | Yes | Yes | Yes | Yes | Yes | Yes | Yes | Yes | Unclear | Unclear | Yes |
| Hult, 2010 | Yes | Yes | Yes | Yes | Yes | Unclear | Yes | Yes | Unclear | Unclear | Yes |
| Kroska, 2018 | Yes | Yes | Yes | Yes | Yes | Yes | Yes | Yes | No | Unclear | Yes |
| Lei, 2016 | Yes | Yes | Yes | Yes | Yes | Yes | Yes | Yes | No | Unclear | Yes |
| Liu, 2016 | Yes | Yes | Yes | Yes | Yes | Yes | Yes | Yes | No | Unclear | Yes |
| Lumey, 2012 | Yes | Yes | Yes | Yes | Yes | Yes | Yes | Yes | Yes | Yes | Yes |
| Mazumder, 2010 | Yes | Yes | Yes | Yes | Yes | Yes | Yes | Yes | Unclear | Unclear | Yes |
| Mryskyla, 2013 | Yes | Yes | Yes | Yes | Yes | Yes | Yes | Yes | Unclear | Unclear | Yes |

| | | | | | | | | | | | |
|---|---|---|---|---|---|---|---|---|---|---|---|
| Painter, 2006 | Yes | Yes | Yes | Yes | Yes | Yes | Yes | Yes | Yes | Yes | Yes |
| Ravelli, 1999 | Yes | Yes | Yes | Yes | Yes | Yes | Yes | Yes | Yes | Yes | Yes |
| Roseboom, 2001 | Yes | Yes | Yes | Yes | Yes | Yes | Unclear | Yes | Yes | Yes | Yes |
| Schrier, 2010 | Yes | Yes | Yes | Yes | Yes | Yes | Yes | Yes | Unclear | Unclear | Yes |
| Sotomayer, 2013 | Yes | Yes | Yes | Yes | Yes | Unclear | Yes | Yes | Unclear | Unclear | Yes |
| Stein, 2006 | Yes | Yes | Yes | Yes | Yes | Yes | Yes | Yes | Yes | Yes | Yes |
| **Adult** | | | | | | | | | | | |
| Baum, 2019 | Yes | Yes | Yes | Yes | Yes | | Yes | Yes | Yes | Yes | Yes |
| Brackbill, 2006 | Yes | Yes | Yes | Yes | Yes | Yes | Yes | Yes | Unclear | Yes | Yes |
| Dirkzwager, 2007 | Yes | Yes | Yes | Yes | Yes | Yes | Yes | Yes | Unclear | Unclear | Yes |
| Fonseca, 2009 | Yes | Yes | Yes | Yes | Yes | No | Yes | Yes | Yes | Not applicable | Yes |
| Gautam, 2009 | Yes | Yes | Yes | Yes | No | Yes | Yes | Yes | Unclear | Unclear | Yes |
| Huizink, 2006 | Yes | Yes | Yes | Yes | Yes | No | Yes | Yes | Yes | Yes | Yes |
| Jiao, 2012 | Yes | Yes | Yes | Yes | No | Yes | Yes | Yes | Unclear | Unclear | Yes |
| Jordan, 2011 (A) | Yes | Yes | Yes | Yes | Yes | Yes | Yes | Yes | No | No | Yes |
| Jordan, 2011 (B) | Yes | Yes | Yes | Yes | Yes | Yes | Yes | Yes | Unclear | Yes | Yes |
| Jordan, 2013 | Yes | Yes | Yes | Yes | Yes | Yes | Yes | Yes | Unclear | Yes | Yes |
| Joseph, 2014 | Yes | Yes | Yes | Yes | Yes | Yes | Yes | Yes | Unclear | Unclear | Yes |
| Lawrence, 2019 | Yes | Yes | Yes | Yes | Yes | No | Yes | Yes | Unclear | Not applicable | Yes |
| Moscona, 2019 | Yes | Yes | Yes | Yes | No | Unclear | Yes | Yes | Unclear | Unclear | Yes |
| Ng, 2011 | Yes | Yes | Yes | Yes | Yes | No | Yes | Yes | Unclear | Unclear | Yes |
| Peters, 2013 | Yes | Yes | Yes | No | No | No | Yes | Yes | Unclear | Unclear | Yes |
| Rey, 2007 | Yes | Yes | Yes | Yes | Yes | Yes | Yes | Yes | Unclear | Unclear | Yes |

| | | | | | | | | | | |
|---|---|---|---|---|---|---|---|---|---|---|
| Thethi, 2010 | Yes | Yes | Yes | No | No | No | Yes | Yes | Yes | Unclear | Yes |
| Trasande, 2018 | Yes | Yes | Yes | Yes | Yes | Yes | Yes | Yes | Unclear | Not applicable | Yes |
| Vanasse, 2016 | Yes | Yes | Yes | Yes | Yes | Yes | Yes | Yes | Unclear | Not applicable | |
| Yu, 2018 | Yes | Yes | Yes | Yes | Yes | Yes | Yes | Yes | No | No | Yes |

**Table A3.** Critical appraisal using Joanna Briggs Institute (JBI) checklist for cross-sectional studies by exposure period (n=7)

| **Author, Year** | **Were the criteria for inclusion in the sample clearly defined?** | **Were the study subjects and the setting described in detail?** | **Was the exposure measured in a valid and reliable way?** | **Were objective, standard criteria used for measurement of the condition?** | **Were confounding factors identified?** | **Were strategies to deal with confounding factors stated?** | **Were the outcomes measured in a valid and reliable way?** | **Was appropriate statistical analysis used?** |
|---|---|---|---|---|---|---|---|---|
| **Perinatal** | | | | | | | | |
| Bercovich, 2014 | Yes | Yes | Yes | Yes | Yes | Yes | No | Yes |
| Oni, 2015 | Yes | Yes | Yes | Yes | Yes | Yes | Yes | Yes |
| **Adult** | | | | | | | | |
| An, 2015 | Yes | Yes | Yes | Yes | No | No | Yes | Yes |
| Bich, 2011 | No | Yes | Yes | Yes | No | No | No | Yes |
| Husarewycz, 2014 | Yes | Yes | Yes | Yes | Yes | Yes | Yes | Yes |
| Katratzias, 2015 | Yes | Yes | Yes | Yes | Yes | Yes | Yes | Yes |
| Koroma, 2019 | No | No | Yes | Yes | No | No | Yes | Yes |

**Table A4.** Critical appraisal using Joanna Briggs Institute (JBI) checklist for quasi-experimental studies by exposure period (n=10)

| Author, Year | Is it clear in the study what is the 'cause' and what is the 'effect' (i.e. there is no confusion about which variable comes first)? | Were the participants included in any comparisons similar? | Were the participants included in any comparisons receiving similar treatment/care, other than the exposure or intervention of interest? | Was there a control group? | Were there multiple measurements of the outcome both pre and post the intervention/exposure? | Was follow up complete and if not, were differences between groups in terms of their follow up adequately described and analyzed? | Were the outcomes of participants included in any comparisons measured in the same way? | Were outcomes measured in a reliable way? | Was appropriate statistical analysis used? |
|---|---|---|---|---|---|---|---|---|---|
| **Perinatal** | | | | | | | | | |
| Xiao, 2019 | Yes | Unclear | Not applicable | Yes | No | Yes | Yes | Yes | Yes |
| **Adult** | | | | | | | | | |
| Becquart, 2018 | Yes | Unclear | Not applicable | No | No | Yes | Yes | Yes | Yes |
| Gerin, 2005 | Yes | Yes | Not applicable | Yes | Yes | Yes | Yes | Yes | Yes |
| Hendrickson, 1996 | Yes | Unclear | Not applicable | No | Yes | Yes | Yes | Yes | Yes |
| Kim, 2017 | Yes | Unclear | Not applicable | No | Yes | Yes | Yes | Yes | Yes |
| Kong, 2019 | Yes | Unclear | Not applicable | Yes | No | Yes | Yes | Yes | Yes |
| Lin, 2010 | Yes | Unclear | Not applicable | Yes | No | Yes | Yes | Yes | Yes |
| McKinney, 2011 | Yes | Unclear | Not applicable | No | Yes | Yes | Yes | Yes | Yes |
| Nagayoshi, 2015 | Yes | Unclear | Not applicable | Yes | No | Yes | Yes | Yes | Yes |
| Silva-Palacios, 2015 | Yes | Unclear | Not applicable | No | No | Yes | Yes | Yes | Yes |
